# Patients with rheumatoid arthritis with moderate or high disease activity benefit from multidisciplinary rehabilitation: a nested case-control study

**DOI:** 10.1101/2021.04.28.21256249

**Authors:** Bernardo M Cunha, Bruno SA Ferreira, Camila SM Barros, Jesiniana R Silva, Juliana B Kauer, Larissa A Moreira, Andreia Gushikem, GIRE - Grupo de Investigação em REabilitação REumatológica

**Author notes:** Corresponding author: Bernardo Matos da Cunha, MD, PhD., Address: SMHS 501 bloco A, Brasília-DF, Brazil, ZIP code: 70335-901. This research did not receive any specific grant from funding agencies in the public, commercial, or not-for-profit sectors.

## Abstract

**Background:** rheumatologists recognize the importance of rehabilitation in patients with rheumatoid arthritis (RA), but they are not confident if patients with significant disease activity would benefit from it.

**Objective:** To verify if rheumatoid arthritis patients with moderate to severe inflammatory activity (MHA) improve functional capacity (FC) after a comprehensive rehabilitation program.

**Methods:** Nested case-control study. RA patients who completed a rehabilitation program between June 2014 and December 2017 were included. The interventions were prescribed according to the rehabilitation team’s discretion. FC was assessed with Health Assessment Questionnaire Disability Index (HAQ) and compared between before and after interventions. The group which improved at least 50% in CDAI was compared to the group which achieved <50%.

**Results:** We included 46 patients with complete HAQ and baseline CDAI data, with a mean age of 53.6 years and mean disease duration of 11.8 years. HAQ and CDAI improved on average 0.481 (± 0.500) and 14.2 (± 16.7), respectively. Patients who improved CDAI tended to have a greater mean HAQ difference (0.6 vs. 0.3; p = 0.058). Conversely, patients who did not improve disease activity had a HAQ reduction of 0.3 (± 0.4). Post-hoc analysis was performed on the group of 9 patients with baseline CDAI ≤10. A mean baseline CDAI of 5.2 and a mean HAQ difference of 0.319 (0.079; 0.56; p = 0.016) were found.

**Conclusions:** After rehabilitation, RA patients with sustained MHA improved FC similarly to patients with baseline mild activity or remission. Thus, patients with RA and MHA may benefit from rehabilitation concurrently with drug treatment. This study suggests that the range of improvement in FC with rehabilitation appears to have an additive effect to the drug therapy.

## Introduction

Rheumatoid arthritis (RA) is a prevalent joint disease, affecting 0.8% of the XXX population ^1^. RA can lead to disabling joint pain and severe sequelae. Patients with RA have reduced functional capacity compared to the general population ^2^.

Among other effects, adequate treatment with disease-modifying drugs (DMARDs) improves the functional capacity (FC) of patients with RA ^3,4^. Additionally, rehabilitation strategies have additive beneficial effects in this population and are safe and well tolerated ^5^. But beyond adequate pharmacologic treatment, favorable RA rehabilitation outcomes are associated with degree of functional disability, level of pain or fatigue, decreased psychological well-being and better health perception ^6,7^.

The *Grupo de Investigação em Reabilitação Reumatológica* of Hospital XXX intends to dynamically and prospectively evaluate rehabilitation interventions in patients with inflammatory joint diseases, including RA. The Rehabilitation Program in Rheumatology of XXX provides a multidisciplinary approach, with the simultaneous participation of physicians, physiotherapists, occupational therapists, nurses, social workers, psychologists and physical educators. The program started in August 2014 and offers multimodal outpatient interventions. The predominant clientele of Hospital XXX is the population from the metropolitan region of XXX.

Several barriers exist for patient compliance to exercise-based rehabilitation programs, such as fear of worsening pain, fatigue, stiffness, lack of mobility and lack of specific programs for RA patients ^8^. Although health professionals, including rheumatologists, recognize the importance of physical activity in this group of patients ^9^, they are not confident how to promote patients’ engagement in physical activity and do not address these issues in the first consultations. If patients are in moderate to high inflammatory disease activity, health professionals are often afraid that patients might worsen their sequelae and because they are not confident that they will benefit from the rehabilitation activities.

The objective of this study is to compare improvement in FC after a comprehensive multidisciplinary rehabilitation program of adults with RA with baseline MHA to adults with baseline RLDA.

## Materials and Methods

### Design

Retrospective cohort study from the prospective Sarar 2 cohort. The project was submitted to the XXX (national ethics committee site) with CAAE 71775317.0.0000.0022 and was approved by the XXX Ethics in Research Committee. The work described has been carried out in accordance with The Code of Ethics of the World Medical Association for experiments involving humans.

### Inclusion criteria

All patients aged ≥18 years with rheumatoid arthritis, defined according to the 2010 ACR/EULAR criteria ^10^, who completed the rehabilitation program between June 2014 and December 2017 and had complete Health Assessment Questionnaire Disability Index (HAQ) and baseline Clinical Disease Activity Index (CDAI) registered data.

### Exclusion criteria

Patients with juvenile idiopathic arthritis and patients who underwent surgery during the evaluation period were excluded.

### Assessments

The assessments were performed in the context of patient care in the rehabilitation in rheumatology outpatient clinic. This clinic has the permanent participation of three rheumatologists, a physiotherapist and a nurse. Other healthcare professionals participate in the patient care on demand.

The baseline demographic characteristics, clinical and laboratory data from patients were extracted from their medical records: age, gender, address, level of schooling, disease duration, rheumatoid factor, ACPA, C-reactive protein, hemosedimentation rate, DMARDs and rehabilitation modalities. The FC was evaluated with the HAQ validated for Brazilian Portuguese ^11^. HAQ can be scored as 0 to 3, meaning better FC in lower scores. The disease activity was assessed with the CDAI ^12^ CDAI can be scored as 0 to 40, meaning lower disease activity in lower scores. We registered CDAI total scores and each of its components: number of swollen and tender joints, patient and physician global assessment. Patients collected laboratory exams as required for their rheumatologists in the beginning of rehabilitation.

### Interventions

The interventions were individually planned at the discretion of the rehabilitation team. They included drug treatment, joint injections, usual or water dynamic exercises, orthoses, insoles, assistive devices, guidance on joint protection and energy conservation measures (functional training), educational activities, body perception and group psychological interventions. They were planned in a case-by-case analysis. The drug treatment included conventional synthetic, target-specific and biological DMARDs (table 1). Multiple rehabilitation interventions were performed (table 2). Pharmacologic therapy could be modified according to rheumatologists’ evaluation during the rehabilitation period.

**Table 1.**
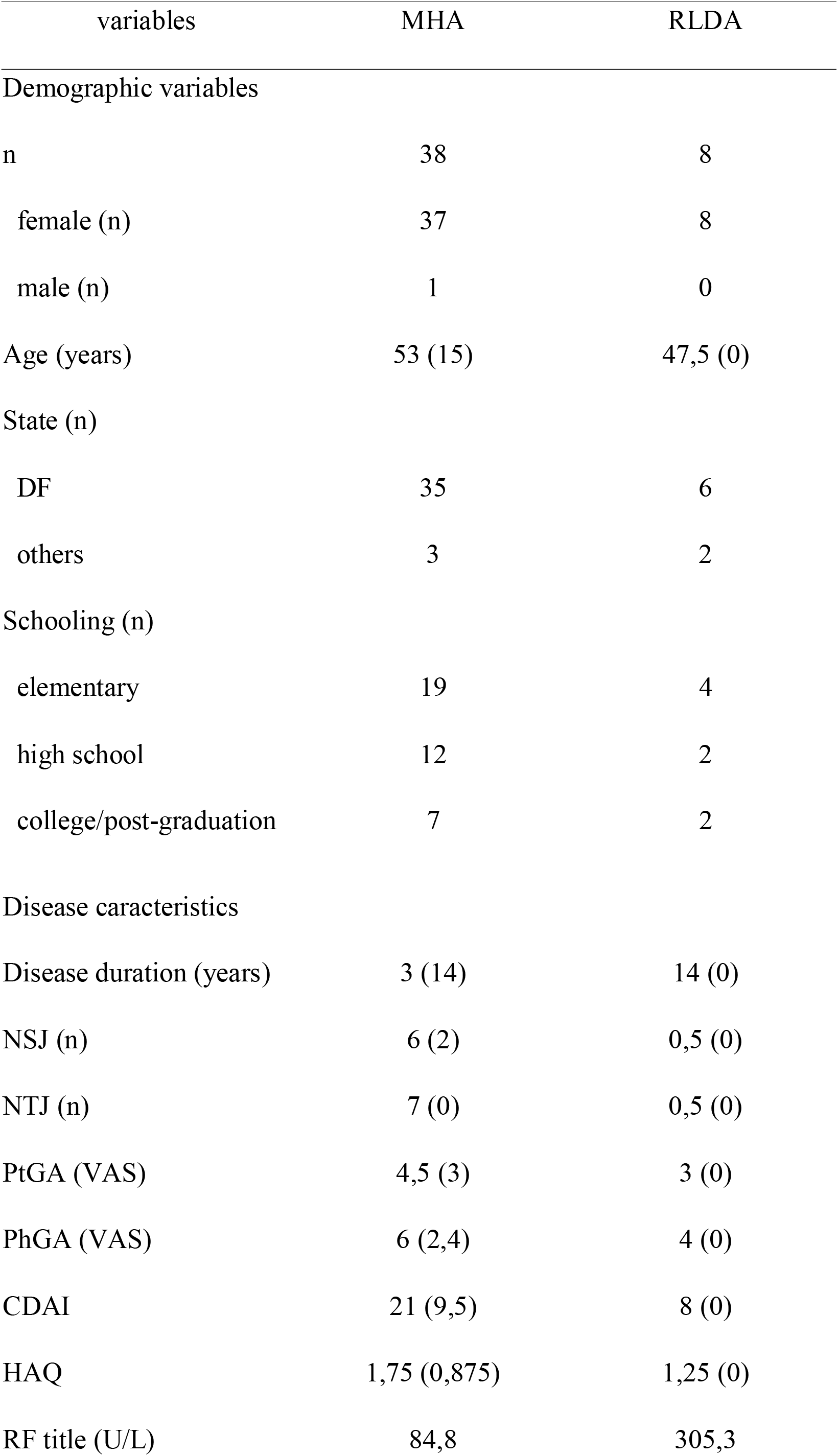
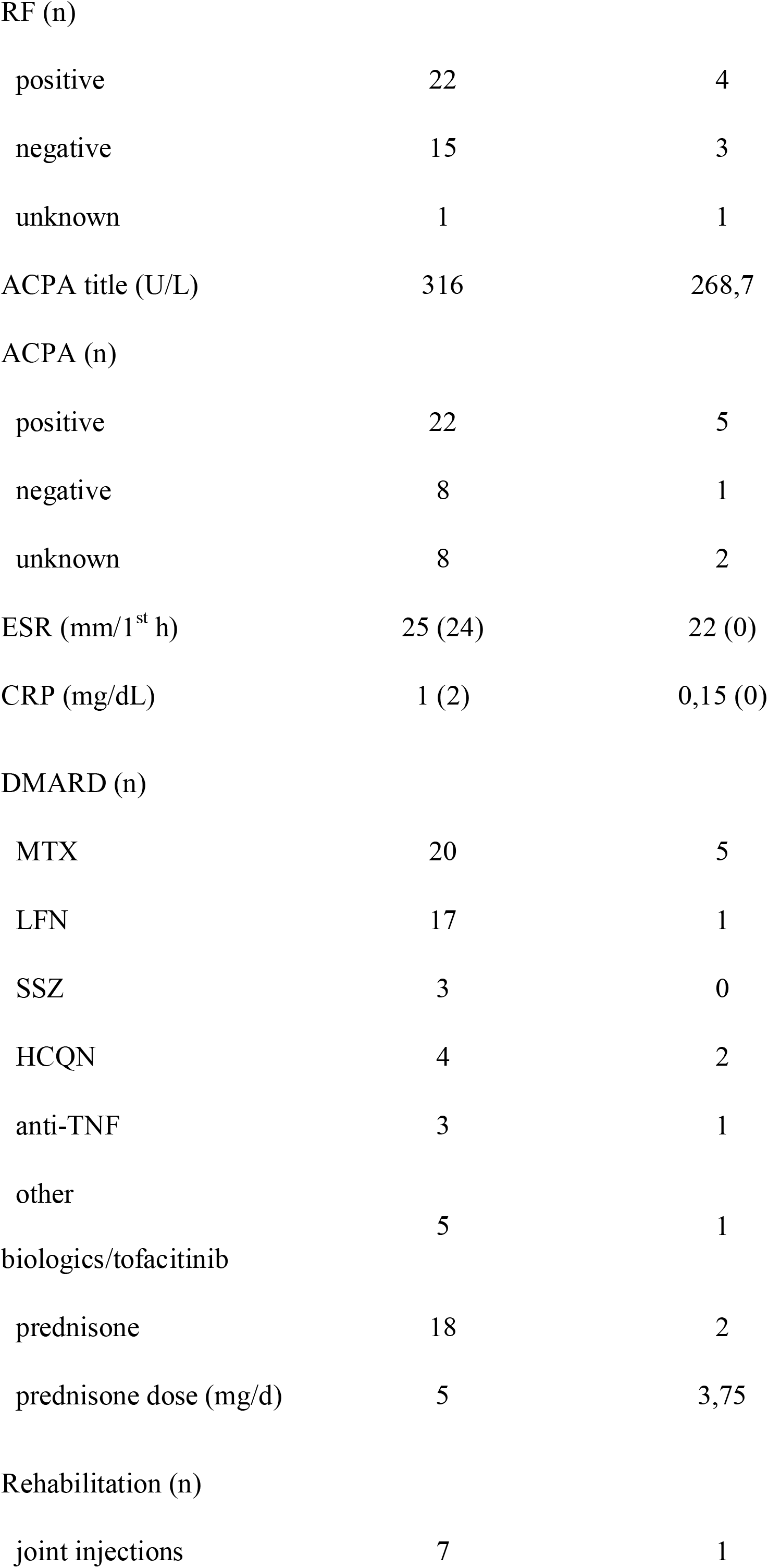
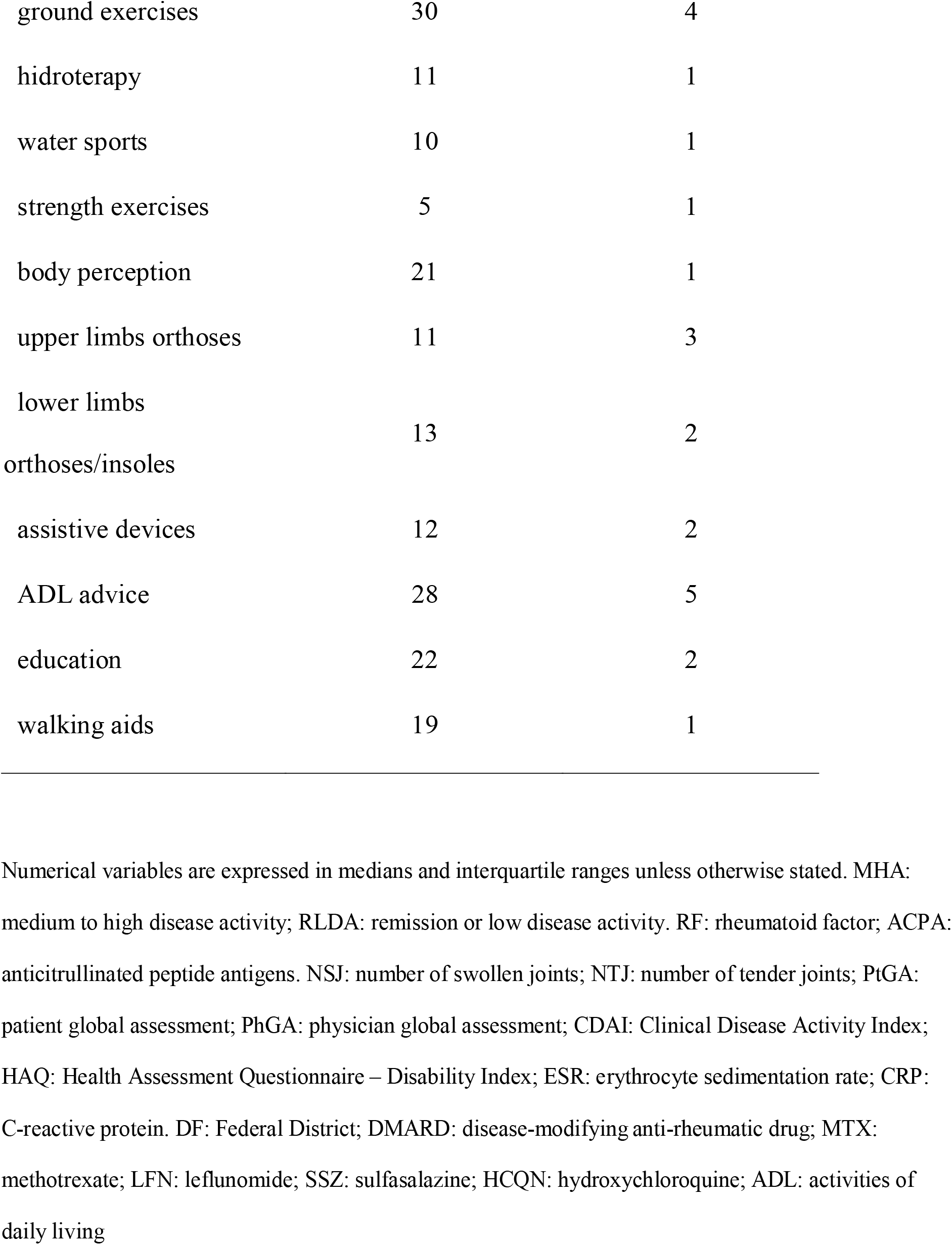
Baseline caracteristics of patients

**Table 2.** Baseline characteristics of patients with MHA who improved the Health Assessment Questionnaire-Disability Index vs. those who did not improve, at the end of the rehabilitation programme.

| Variables | HAQ not improved | HAQ improved | p |
| --- | --- | --- | --- |
| Demographic |  |  |  |
| n | 11 | 27 |  |
| Age (years) | 54 (7,4) | 57 (12,6) | 1 |
| Schooling (n) |  |  |  |
| elementary | 6 | 12 |  |
| high school | 2 | 4 | 0,949 |
| college/post-graduation | 2 | 5 |  |
| Disease characteristics |  |  |  |
| disease duration (years) | 7,8 (11,2) | 15,4 (14,5) | 0,682 |
| CDAI | 29,3 (14,9) | 21,6 (12,3) | 0,924 |
| HAQ | 1,33 (0,7) | 1,75 (0,6) | 0,89 |
| ESR (mm 1<sup>st</sup>h) | 24 (14,5) | 36,5 (18,2) | 0,331 |
| CRP (mg/dL) | 1,1 (0,9) | 1,4 (1,5) | 0,45 |
| RF positivity (n) | 7/19* | 12/19* | 1 |
| RF titre (U/L) | 371,4 (687,2) | 234,1 (262,3) | 0,977 |
| ACPA positivity (n) | 6/15* | 9/15* | 0,613 |
| ACPA titre (U/L) | 227,6 (204,5) | 399,8 (341,7) | 0,392 |
| DMARD (n) |  |  |  |
| MTX | 6 | 9 | 0,458 |
| LFN | 5 | 3 | 0,685 |
| SSZ | 0 | 3 | 0,533 |
| HCQN | 0 | 2 | 1 |
| anti-TNF | 1 | 6 | 0,379 |
| other biologics/tofacitinib | 1 | 4 |  |
| prednisone | 4 | 7 | 1 |
| Rehabilitation (n) |  |  |  |
| joint injections | 2 | 4 | 1 |
| land exercises | 8 | 14 | 0,677 |
| hidrotherapy | 4 | 5 | 0,652 |
| water sports | 2 | 8 | 0,677 |
| strength exercises | 1 | 3 | 1 |
| body perception | 0 | 4 | 0,277 |
| upper limbs orthoses | 5 | 7 | 0,447 |
| lower limbs orthoses/insoles | 4 | 6 | 0,685 |
| assistive devices | 3 | 7 | 1 |
| functional training | 6 | 15 | 0,685 |
| education activities | 1 | 5 | 0,634 |
| walking aids | 4 | 6 | 0,685 |

Numerical variables are expressed in means and standard deviations. CDAI: Clinical Disease Activity Index. HAQ: Health Assessment Questionnaire – Disability Index. ESR: erythrocyte sedimentation rate. CRP: C-reactive protein. RF: rheumatoid factor. ACPA: anticitrullinated peptide antigens. MTX: methotrexate. LFN: leflunomide. SSZ: sulfasalazine. HCQN: hydroxychloroquine. ESR: erythrocyte sedimentation rate. CRP: C-reactive protein.. \* Patients with antibody positivity.

### Statistics

The patients were divided into two groups, according to the baseline CDAI. The cut-off point was set at 10 to split patients in 2 groups: RLDA and MHA. Descriptive statistics was used for baseline characteristics. Included patients were compared with excluded patients, to assess report bias.

A clinically significant improvement was defined for HAQ as a minimum difference of -0.220 ^13^ and for CDAI as a minimum improvement of 50% ^14^. Mean results before and after interventions in each group were compared with the paired t-test. The mean differences were calculated, along with their respective confidence intervals.

Within the MHA group, we planned a nested case-control analysis. The differences between baseline and follow-up HAQ of patients who improved vs. those who did not improve the CDAI after the interventions were compared with the independent samples t-test, as we wanted to know if HAQ improvement could be attributed only to drug therapy.

In all analyses, a 95% significance level was assumed. Analyses were performed in SPSS version 21.

## Results

We included 38 patients with MHA and 8 patients with RLDA with complete HAQ and baseline CDAI data, from a sample of 78 patients. The other 32 patients were excluded due to incomplete data. The population had a mean age of 53.6 (±11.9) years and mean duration of 11.8 (±13.3) years of illness. The baseline characteristics are in table 1. Groups were similar at baseline (table 1). Baseline CDAI and RF title trended to be greater in patients who did not improve, as well as a greater time from diagnosis, baseline HAQ and ACPA title in patients who improved, but without statistical significance (table 2).

In the MHA group, the HAQ and CDAI improved on average 0.481 (0.322;0.640, p<0.001) and 14.4 (8.7;20.2, p<0.001) respectively. All CDAI domains improved (supplementary data). In the RLDA group, the HAQ improved 0.319 (0.079;0.560, p=0.016) on average and, as expected, the CDAI did not improve significantly.

Patients with MHA who improved their CDAI were compared with those who did not improve. In the MHA group, the patients who improved their CDAI during the rehabilitation period had a trend to a greater mean difference of HAQ compared to the patients with MHA who did not improve their CDAI (0.60 vs. 0.31, difference 0.29 (−0.6;0); p = 0.058).

## Discussion

RA patients with baseline MHA who participated in a multidisciplinary rehabilitation program concurrent with drug treatment significantly improved FC. Patients with sustained MHA, i.e., who did not improve the CDAI throughout the study, had an improvement in FC at a similar magnitude of patients in RLDA (0,310 vs. 0,319, respectively). In contrast, patients with baseline MHA who improved CDAI presented an additional benefit in HAQ (0,600). Therefore, our data suggest that DMARD treatment and rehabilitation of patients with MHA improve the HAQ by approximately 0.3 each, in an additive way. Our rehabilitation center has a structure that allows for the possibility of multiple simultaneous interventions, so it may be difficult to isolate the weight of each one in a “real life” setting.

Our study documented the improvement in functional capacity in patients with different levels of disease activity in a “real world” setting, although some randomized controlled trials had published evidence of benefits in patients with mixed levels of disease activity ^15–18^. Recently, Szewczyk et al published an article with a similar design to this study, but with some important differences ^19^. First, they split the disease activity groups in a different way: they included remission to moderate disease activity patients within a group and high disease activity patients in the other, while we included moderate to severe patients in the same group and remission to low disease activity patients in the other. Second, their patients’ rehabilitation lasted a smaller amount of time than ours. Third, they did not employ assistive devices, insoles, orthoses, body perception and patient education, while we did not employ electrotherapy, ultrasound, low-frequency magnetic field and laser therapy. Even so, they found a similar improvement in HAQ-DI to our patients with moderate to high disease activity who kept stable along the course of rehabilitation program. Our patients we older, what might account for the greater time to reach the same benefit. Fourth, they did not include patients with modifications in disease activity, but it is not possible to exclude an improvement in DAS-28 as a result of a late effect of drug therapy, since they included patients in a 3-month window of treatment stability and some patients may take 6 months to improve. We compared patients who improved CDAI to the ones who did not improve, to overcome this limitation. To our knowledge, our study was the first to quantify the amount of benefit of each intervention (drug therapy and rehabilitation) while performed simultaneously. Szewczyk et al did not inform time since diagnosis, therefore we could not estimate if the amount of sequelae in their study was dissimilar from ours. Both studies did not inform radiographic scores, but it would be unpractical, since in our cohort the patients were very distinct about their injured joints.

International guidelines suggest that a multidisciplinary treatment should be offered to patients with RA if possible ^18^, but there are still relatively few specialized rehabilitation services for patients with inflammatory joint diseases ^5^. Our study suggested that even patients with MHA improve their FC after rehabilitation. Considering the results of this study, rheumatologists and rehabilitation health professionals should be more confident to offer a rehabilitation program for patients with RA. A multicentric cohort study could be performed for cost-effectiveness analyses of a comprehensive rehabilitation program.

This study has limitations. This study included a relatively small population, but we plan to update it in a regular basis. The association between the presence of joint sequelae and functional capacity was not evaluated due to the complexity of interpreting these data from patients with multiple sequelae in a small population. Psychological aspects and fatigue were not assessed. Concomitant fibromyalgia was not included as a covariate. We did not perform a formal safety evaluation, but no patients had musculoskeletal lesions during rehabilitation.

We plan to perform the following studies in the future: best rehabilitation modalities for each level of disease activity; assessment of concurrent fibromyalgia and psychological aspects, as self-efficacy or prevalence of depressive symptoms.

## Conclusions

Our one-center study suggests that RA patients with baseline MHA may benefit from a multidisciplinary rehabilitation program. Therefore, health professionals should offer a rehabilitation treatment to patients with RA, regardless of their level of disease activity. Patients with RA with MHA may be rehabilitated concomitantly with drug treatment and DMARD treatment and rehabilitation seem to have an additive effect on FC improvement. There is no reason, besides other limitations as cardiovascular disease, not to offer a rehabilitation program to patients with RA, even if they are MHA.

## Data Availability

Data are not available meanwhile.

http://www.sarah.br

## Acknowledgments

We would like to thank Dr. Sergio Henrique Rodolpho Ramalho for helping in translation into English.

## Author Contributions

Bernardo Matos da Cunha: conceptualization, methodology, formal analysis, investigation, data curation, writing-original draft, supervision, project administration.

Bruno Silva de Araújo Ferreira, Camila Sodré Mendes Barros, Jesiniana Rodrigues Silva, Juliana Barnetche Kauer, Larissa Aniceto Moreira and Andreia Gushikem: investigation, writing-review and editing.

## Abbreviations

MHA: moderate or severe disease activity
FC: functional capacity
RLDA: remission or low disease activity

---

**Supplementary table 1.** Functional capacity and disease activity indices before and after the rehabilitation programme.

| Variables | Baseline | Follow-up | Difference (95%CI) | p |
| --- | --- | --- | --- | --- |
| HAQ | 1,59 | 1,08 | 0,51 (0,3-0,71) | **<0,001** |
| CDAI | 22,1 | 9,4 | 12,7 (7,6-17,9) | **<0,001** |
| NSJ (n) | 5 | 2 | 3 (2-4) | **<0,001** |
| NTJ (n) | 9 | 3 | 6 (4-9) | **<0,001** |
| PhGA (VAS) | 4,1 | 1,9 | 2,2 (1,4-3) | **<0,001** |
| PtGA (VAS) | 4,6 | 2,9 | 1,7 (0,7-2,7) | **0,002** |
| ESR (mm/1<sup>a</sup> h) | 32,6 | 23,1 | 9,5 (2,4-16,5) | **0,01** |
| CRP (mg/dL) | 2,1 | 1 | 1,1 (0,3-1,9) | **0,012** |

Numerical variables are expressed in means and standard deviations. HAQ: Health Assessment Questionnaire – Disability Index;
CDAI: Clinical Disease Activity Index. NSJ: Number of Swollen Joints; NTJ: number of tender joints; PhGA: Physician Global
Assessment; PtGA: Patient Global Assessment; ESR: erythrocyte sedimentation rate; CRP: C-reactive protein

